# Acceptance of COVID-19 Vaccine in Pakistan Among Health Care Workers

**DOI:** 10.1101/2021.02.23.21252271

**Authors:** Asmara Malik, Jahanzeb Malik, Uzma Ishaq

## Abstract

**Objective:** Acceptance of the COVID-19 vaccine will impart a pivotal role in eradicating the virus. In Pakistan, health care workers (HCWs) are the first group to receive vaccination. This survey aimed at the level of acceptance to the COVID-19 vaccine and predictors of non-acceptance in HCWs.

**Method:** This was a cross-sectional study design and data were collected through 3rd December 2020 and February 14th, 2021. An English questionnaire was distributed through social media platforms and administration of affiliate hospitals along with snowball sampling for private hospitals.

**Results:** Out of 5,237 responses, 3,679 (70.2%) accepted COVID-19 vaccination and 1,284 (24.5%) wanted to delay until more data was available. Only 5.2% of HCWs rejected being vaccinated. Vaccine acceptance was more in young (76%) and female gender (63.3%) who worked in a tertiary care hospital (51.2%) and were direct patient care providers (61.3%). The reason for rejection in females was doubtful vaccine effectiveness (31.48%) while males rejected due to prior COVID-19 exposure (42.19%) and side effect profile of the vaccine (33.17%). Logistic regression analysis demonstrated age between 51-60 years, female gender, Pashtuns, those working in the specialty of medicine and allied, taking direct care of COVID-19 patients, higher education, and prior OCVID-19 infection as the predictors for acceptance or rejection of COVID-19 vaccine.

**Conclusion:** A high overall acceptance rate was observed among HCWs, favoring a successful nationwide vaccination program in Pakistan.

## Introduction

The novel coronavirus disease 2019 (COVID-19) was declared as a pandemic and an emergency was initiated by World Health Organization (WHO) on 30th January 2020^[1]^. The outbreak revealed itself as clusters of pneumonia of unknown etiology in China^[2]^. A systematic review has outlined a severe form of the disease in 20% of the affected individuals with a mortality rate of 3%^[3]^. As of February 2021, COVID-19 has affected 108 million people worldwide, leading to 2.38 million deaths^[4]^ while Pakistan has reported 560,000 cases and 12,218 deaths^[5]^. Hence, in addition to social distancing measures and personal protective equipment^[6]^, there is a vital need to be vaccinated for COVID-19 to curb the community transmission in Pakistan. Health care workers (HCWs) have an important part in educating the general public about the source of the vaccine and its implications in the coming years^[7]^. In Pakistan, HCWs are being prioritized for an early Chinese-based COVID-19 vaccination program^[8]^. This is being mandated throughout the West, prioritizing high-risk groups, and HCWs being recognized as such. Therefore, it is vital to consider HCW attitudes towards the COVID-19 vaccine as it will lead to a better dissemination of knowledge among the general public.

Given the paucity of data regarding vaccine acceptance in South-East Asia among HCWs, we conducted this survey across multiple healthcare facilities throughout Pakistan to measure the acceptance of COVID-19 vaccine, and to enumerate the reasons underlying vaccine hesitancy among HCWs.

## Methods

### Study Design and Sampling

This study was a cross-sectional design to assess the acceptability of HCWs towards the COVID 19 vaccination program in Pakistan. An English questionnaire was designed on Google Forms from a previous study and modified for HCWs using data capture tools hosted at the Foundation University. No identifying information was presented in the survey and all data were collected anonymously. All rights for sharing the survey questionnaire belongs to the University. The Foundation University Ethical Review Committee approved the study design (Number: FFH/51/DCA/2020).

The questionnaire was distributed on social media platforms and a large coverage was made available by our affiliate institutes in major cities of Pakistan. Snowball sampling was also encouraged for dissemination of the survey questionnaire in primary care and private hospitals. Data were collected through 3rd December 2020 and February 14th, 2021. Informed written consent was taken before final form submission and all adults (age ≥ 18 years) working as HCW were considered eligible to participate in the survey. Incomplete questionnaires were excluded from the final analysis.

### Study Variables and Measures

Demographic data was presented on the first page of the survey questionnaire. It included age, gender, ethnicity, marital status, type of designated work, education, type of medical facility, chronic medical conditions, and prior COVID-19 infection. To assess the acceptance of the COVID-19 vaccine, the respondents were provided the brand name and effectiveness of the vaccine (CanSino Biologics, Tianjin, China; 65.7% effective in preventing symptomatic cases). Respondents were given a question of whether they would accept the above-labeled vaccine as yes or no. As for demographic variables, age was grouped into five categories (18-30, 31 – 40, 41 – 50, 51 – 60, >61 years old); ethnicity was grouped as Punjabi, Sindhi, Balochi, Pashtun, or other to find out which group was interested in vaccinating among HCWs; and designation of work was divided into direct patient care providers (Specialists, general practitioners, medical students, and nursing staff) and non-patient care providers (hospital supporting staff, administration, and pharmacists). Type of education and specialty were broken down into medicine/allied, surgery/allied, diagnostics, or other. Place of work was designated as either a tertiary care hospital, primary care center, or a private clinic.

### Statistical analysis

For analysis of the data, Statistical Package for Social Sciences (SPSS) version 26 (IBM, Armonk, NY, USA.) was used and logistic regression was employed to determine the predictors of HCWs acceptance of COVID-19 vaccine. Continuous variables are presented as mean and standard deviation (SD) and categorical variables as frequency and percentages. Student’s t test was used for continuous variables and chi-square for categorical variables. Univariate analysis was done for unadjusted estimate of odds ratio (OR) and multivariate analysis for adjusted OR. Logistic regression was carried out for predictors of vaccine hesitancy. A p-value of less than 0.05 was considered statistically significant.

## Results

### Respondent Demographics

We received 5,381 responses. One-hundred and forty-four were excluded because of the incomplete survey questionnaire. A total of 5,237 responses were included in this study. More than two-thirds of the respondents were younger than 50 years (76%) and 63.3% were females who had either a bachelor’s (28.2%) or a master’s degree (11.6%). Overall, 51.2% of respondents worked in tertiary care hospitals and 61.3% were direct patient care providers. The majority of the responses were received from Punjab (43.1%) in the specialty of medicine and allied (37.9%). Sixty-three percent had a history of COVID-19 disease before this survey.

### Vaccine Acceptance and Predictors

Out of 5,237 respondents, 3,679 (70.2%) accepted the vaccination process while only 274 (5.2%) rejected it. One-fourth of the HCW’s would review data on COVID-19 vaccine before moving further with the vaccine (24.5%). This is expressed in **Figure 1**. There was a significant association between vaccine acceptance and respondent demographics. Respondent demographic details, chronic medical conditions, and percent acceptance of COVID-19 vaccine are presented in **Table 1** along with age, ethnicity, and gender-adjusted response variables.

**Table 1.**
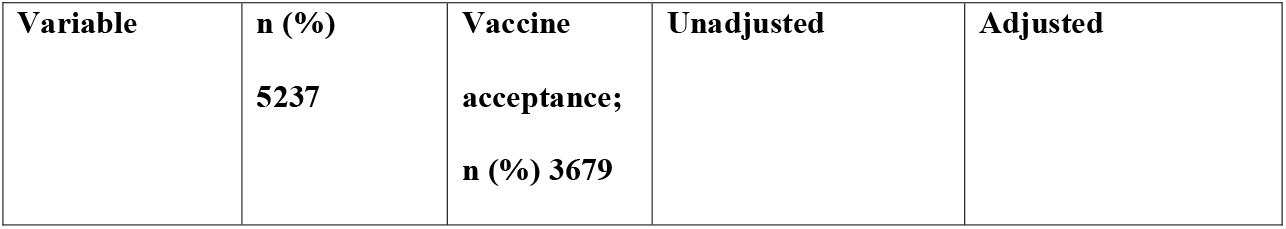
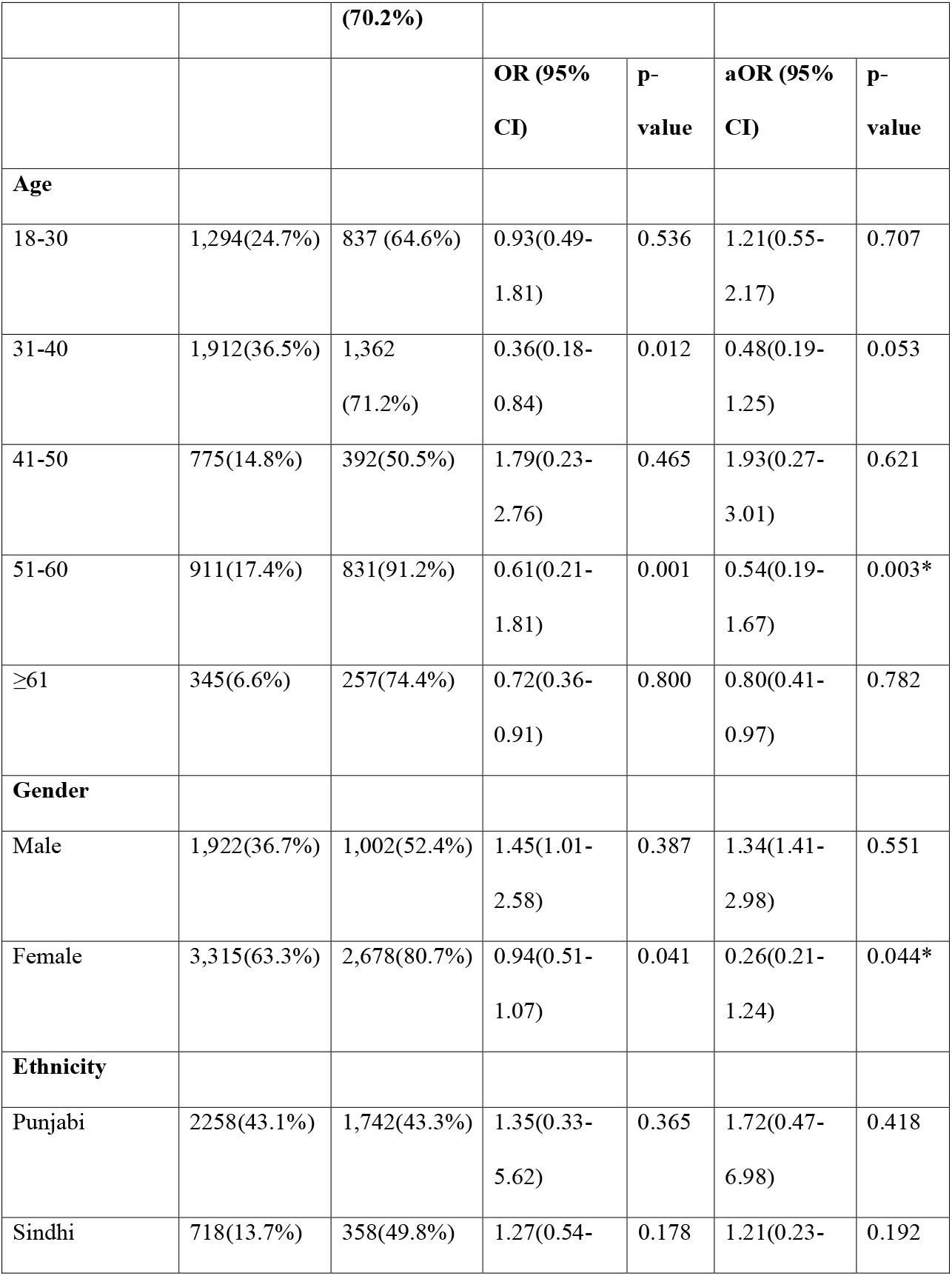
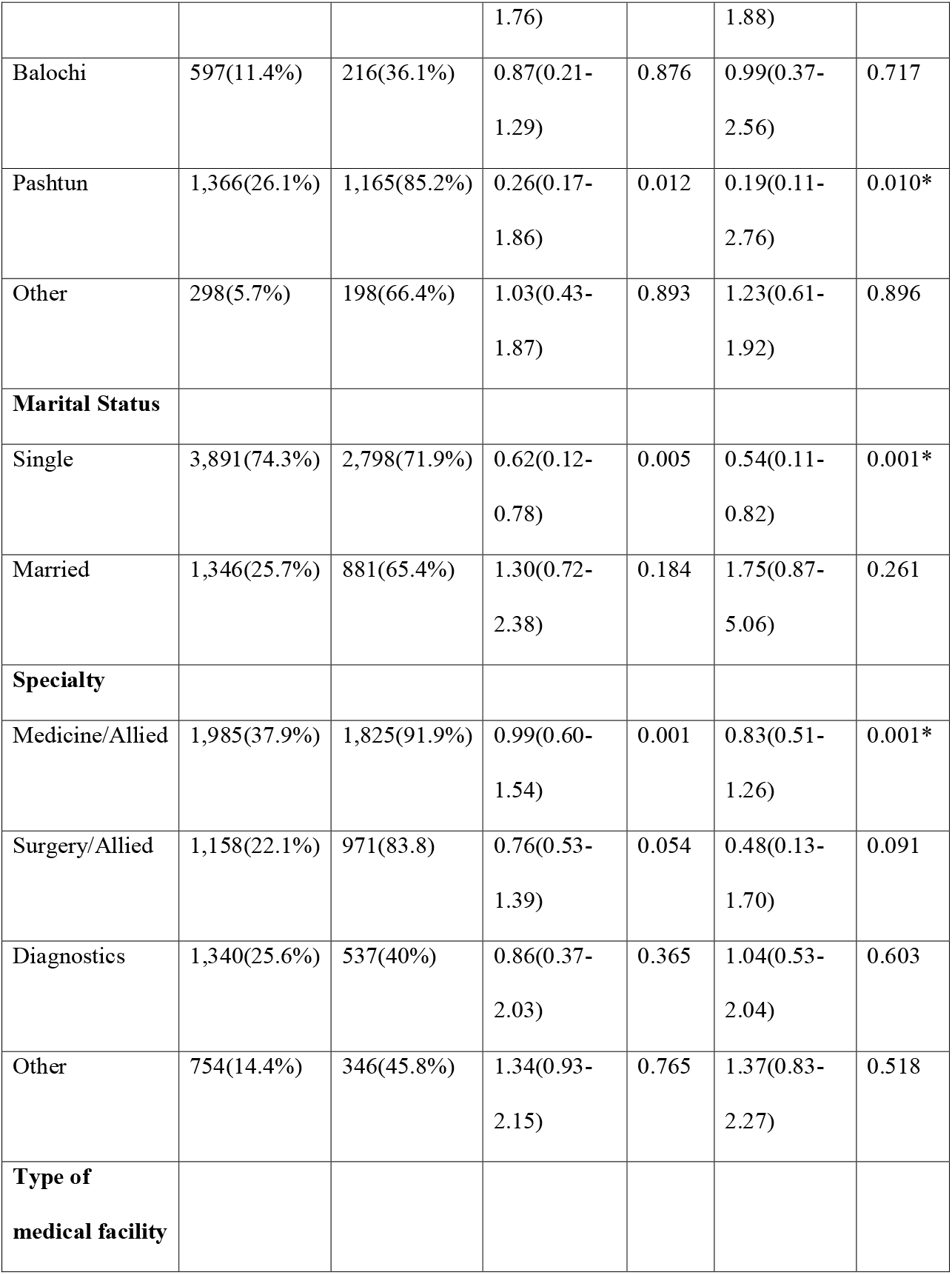
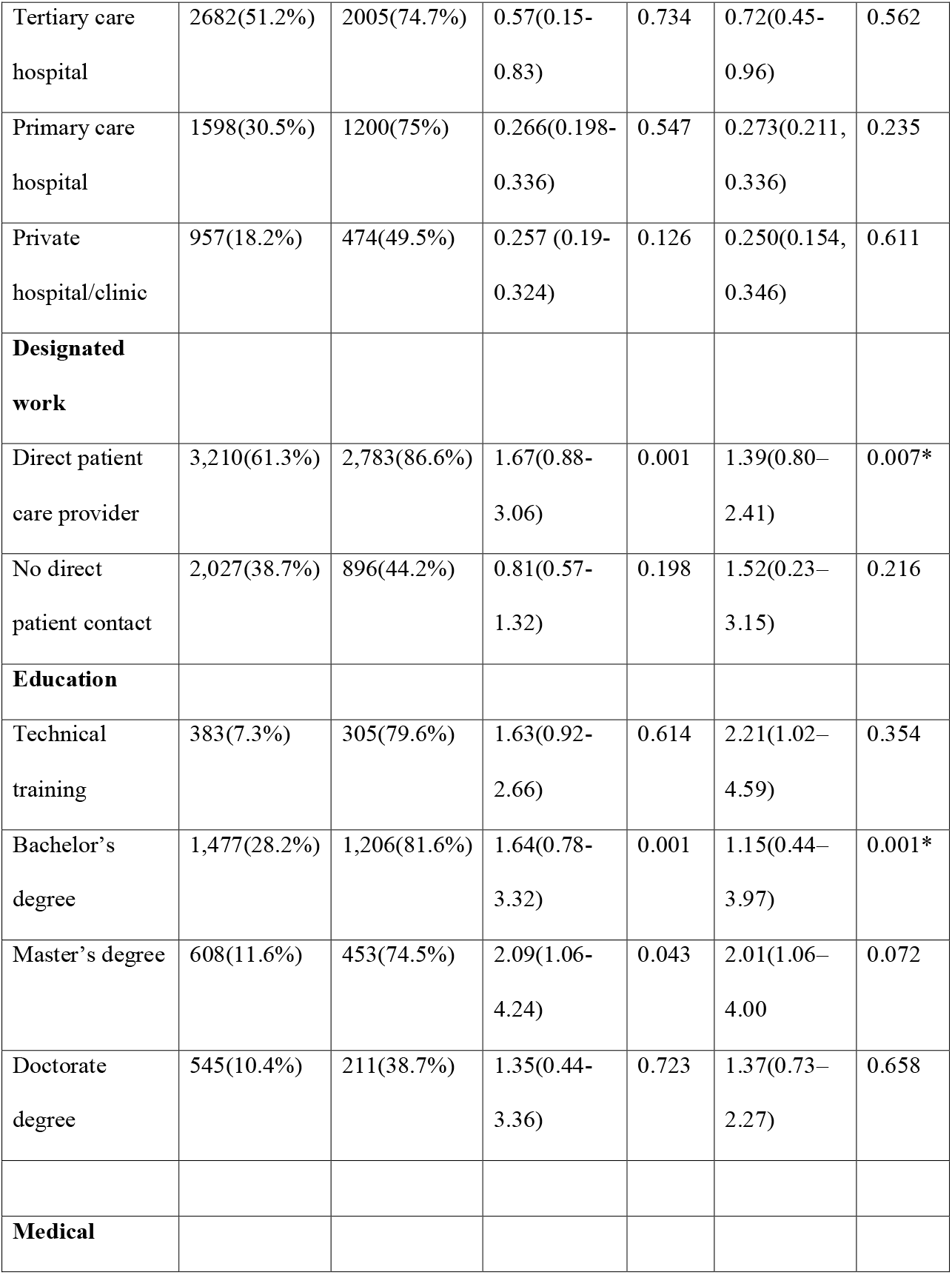
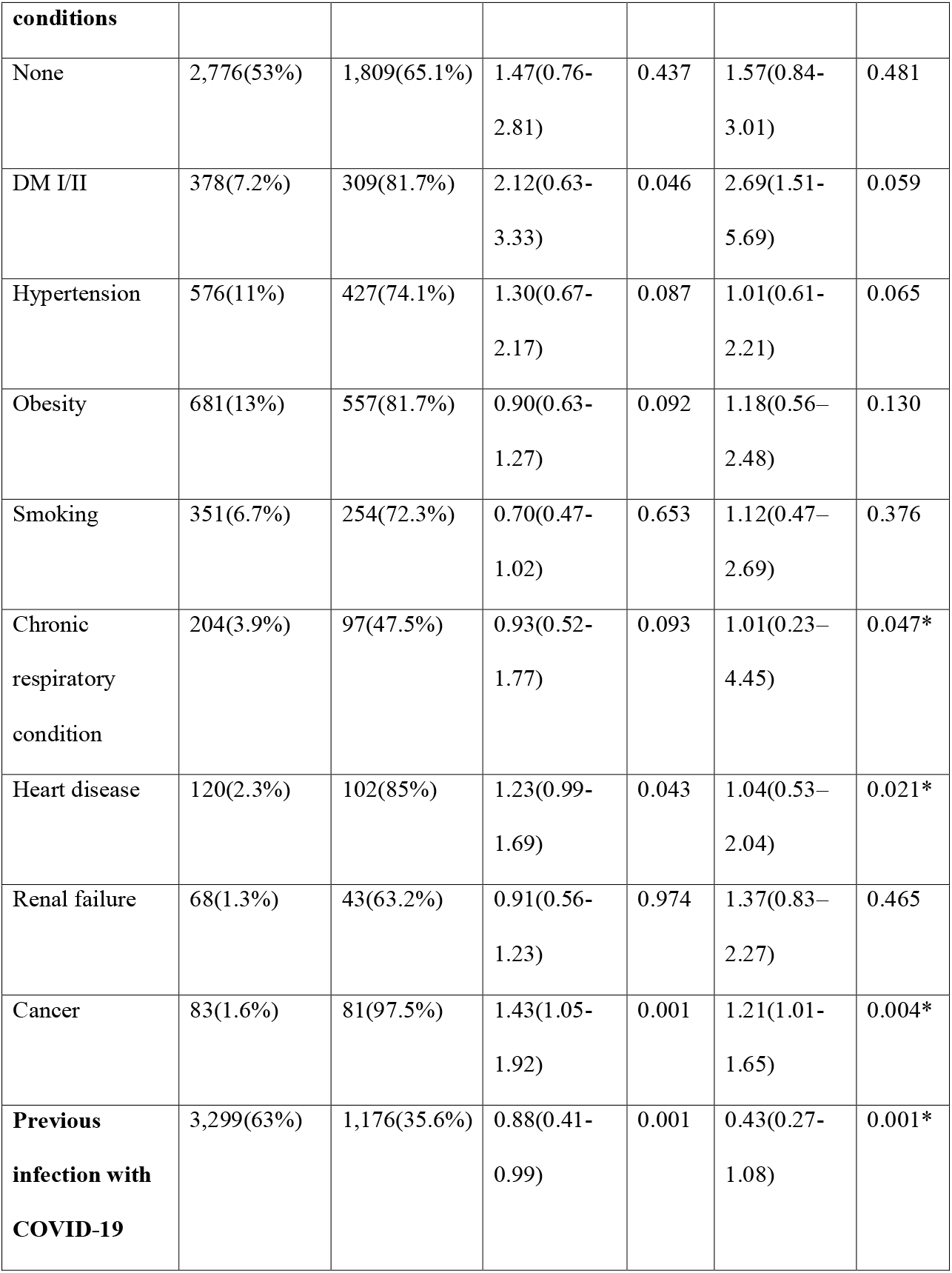
Demographic data and logistic regression analysis demonstrating factors associated with acceptance of a COVID-19 vaccine in health care workers in Pakistan, n=5,237. Variables presented as n (%), adjusted for age, gender, and ethnicity. *p-value <0.05. Diabetes mellitus (DM).

**Figure 1.**
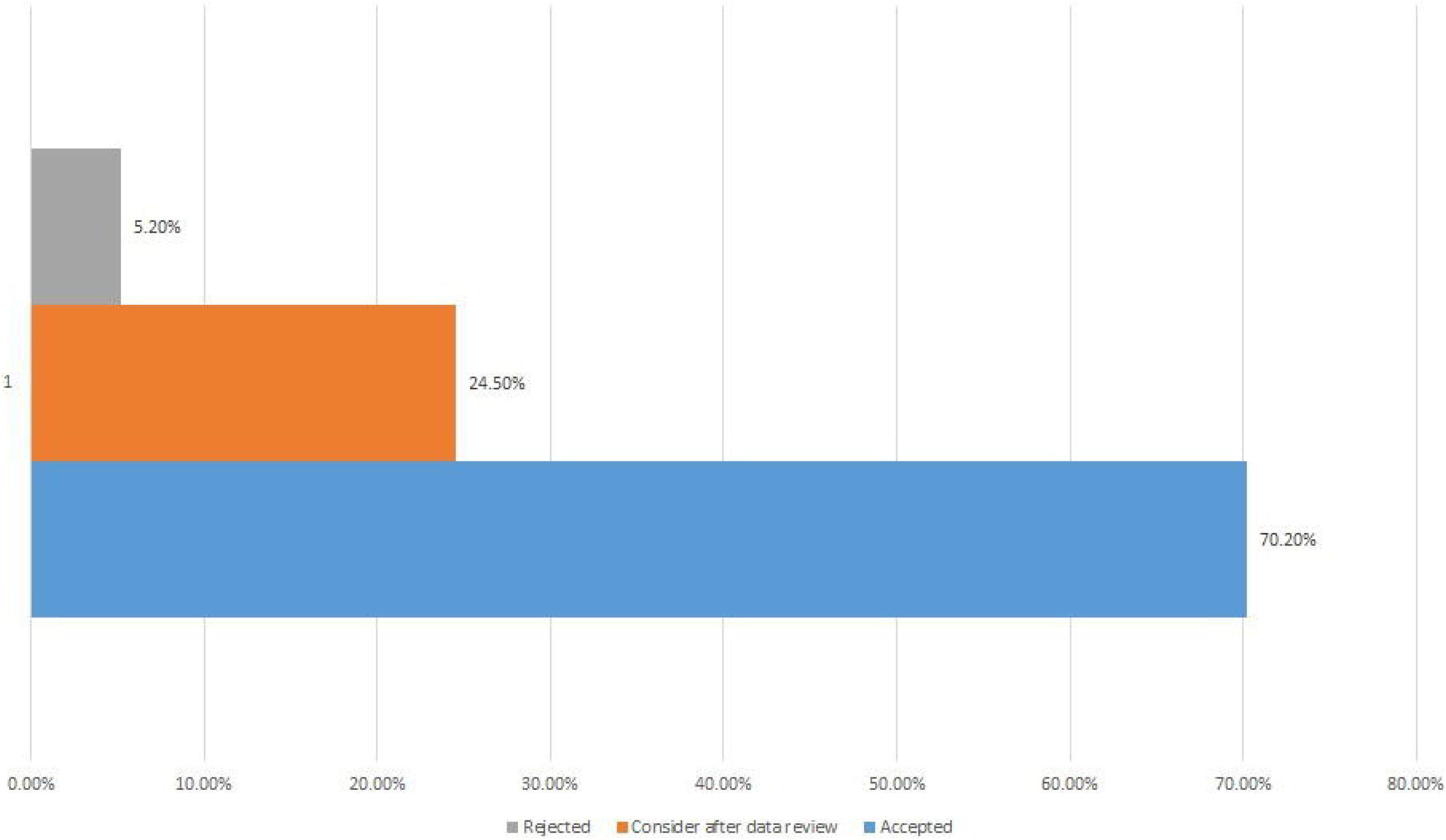
Overall acceptance and rejection rate among HCW’s. Health care workers (HCW)

Acceptance rates of COVID-19 vaccination increased with increasing age. In the 18-30 age group, 64.6% of the respondents accepted the COVID-19 vaccine which increased to 71.2% in 31-40 years and 91.2% in 51-60 years. In >60 years 74.4% accepted for vaccination. Other factors predictive of vaccine acceptance were female gender and single relationship status. The female gender had a higher vaccine acceptance (80.7%) as well as those with a single relationship status (71.9%). A marked difference was seen in the vaccine acceptance among different ethnic groups. Pashtuns (85.2%) had the highest COVID-19 vaccine acceptance while Balochi’s had the lowest acceptance rate (36.1%). Vaccine acceptance varied among various specialties in healthcare. Those working in the specialty of medicine and allied (91.9%), in primary and tertiary healthcare settings (75% and 74.7%) had the highest vaccine acceptance and HCW’s with no direct patient contact had a high refusal rate (55.8%).

Among the two genders, there were different reasons for rejection of the COVID-19 vaccine (**Figure 2**). Females had religious concerns (2.3%) as compared to males (1%) and they were not convinced about the effectiveness of the vaccine (31.48%). The males not willing for the COVID-19 vaccine had prior COVID-19 infection (42.1%) and they were not sure about the side effects of the vaccine (33.1%). Logistic regression analysis demonstrated age between 51-60 years, female gender, Pashtuns, those working in the specialty of medicine and allied, taking direct care of COVID-19 patients, higher education, and prior OCVID-19 infection as the predictors for acceptance or rejection of COVID-19 vaccine (**Figure 3**).

**Figure 2.**
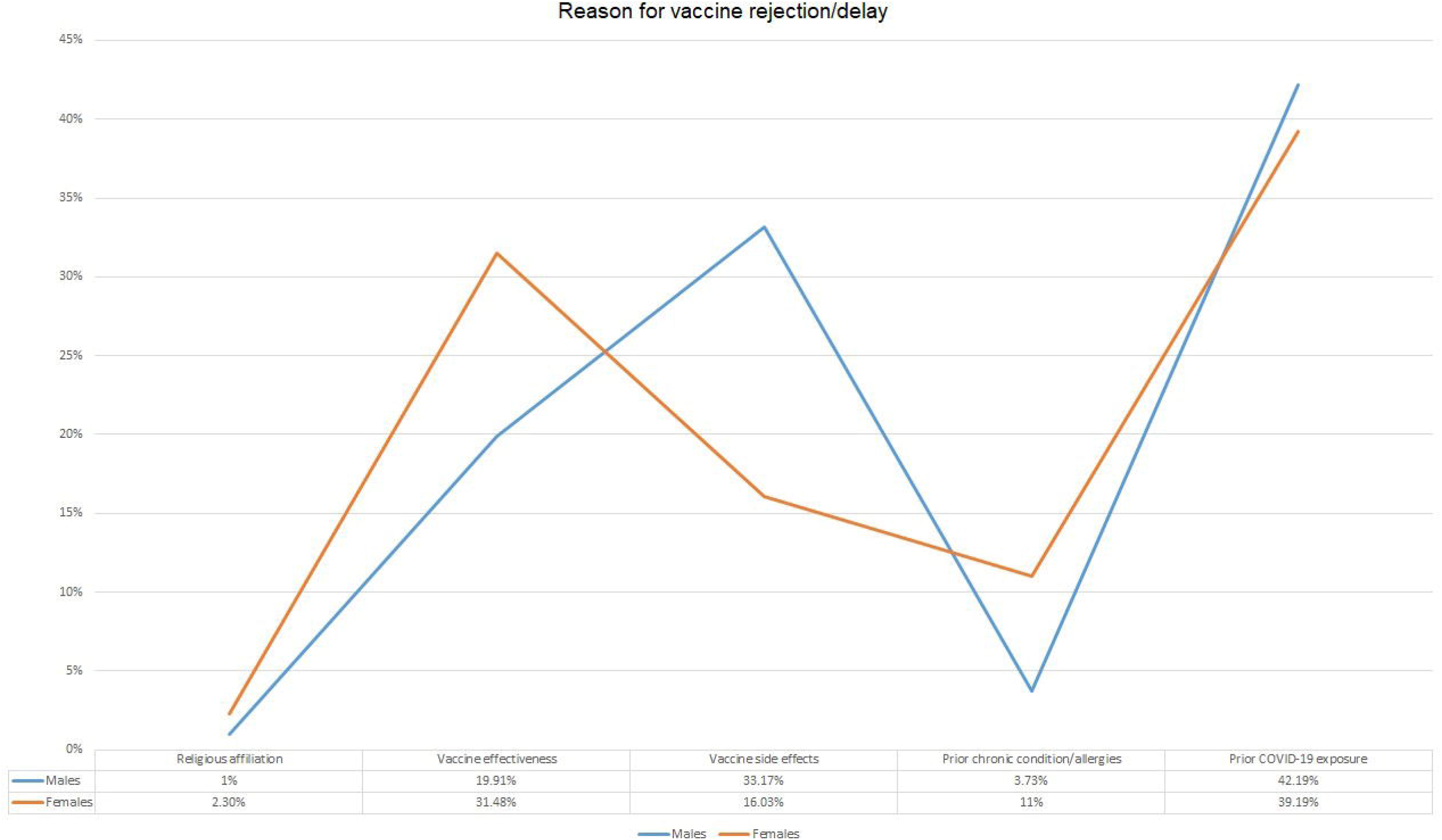
Views among genders regarding non-acceptance of vaccine

**Figure 3.**
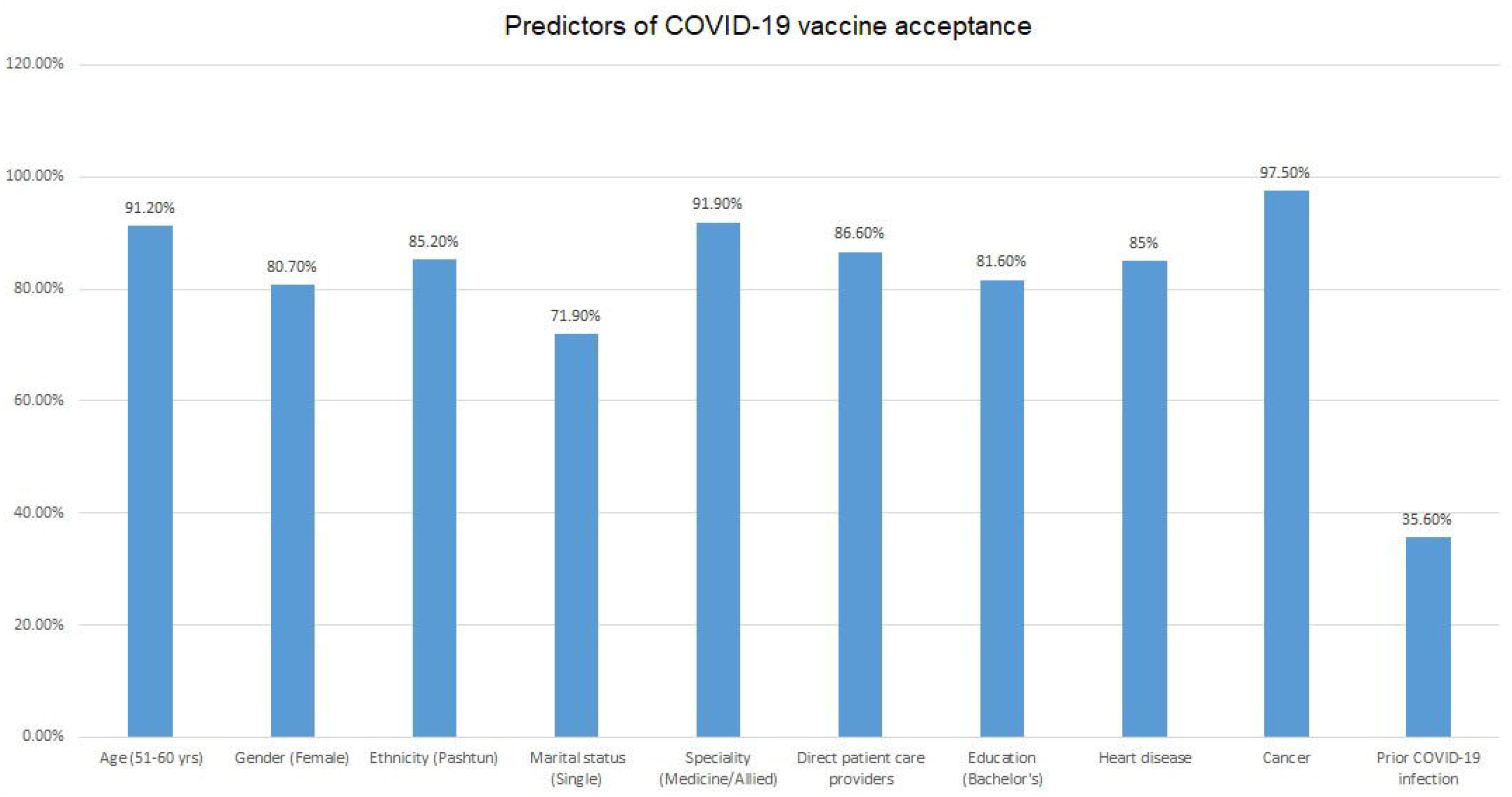
Logistic regression analysis demonstrating predictors for COVID-19 vaccine acceptance

## Discussion

Vaccine hesitancy is an impediment in controlling COVID-19 spread and the hurdles in vaccine uptake among HCWs in South-East Asia is unknown. In our study, most of the HCWs accepted to receive COVID-19 vaccine (70.2%), and only 5.2% rejected it completely. Approximately one-fourth of the respondents wanted more data on the vaccine before going forward with the vaccination process. Taken as a whole, our data suggests that COVID-19 vaccine acceptance among HCWs is higher as compared to general population worldwide. ^[9-14]^ A study presented a forecast model for spread of COVID-19 in Pakistan through an auto-regressive integrated moving average (ARIMA) model. ^[15]^ This forecast can be an underestimation if vaccine is not administered through a sound government campaign.

The incidence of COVID-19 in Pakistani HCWs was consistent with other countries. This may indicate the need for more robust infection control practices in healthcare facilities on a global scale.^[10,20]^ It also reflected a reduced lack of risk perception, especially by male HCWs, who felt decreased need for COVID-19 vaccination in light of a previous infection. This is consistent among HCWs worldwide due to the limited data available about the severity of COVID-19 re-infection and long-term health impairment even after recovery from COVID-19. ^[21,22]^ In contrast to a study of US healthcare workers ^[23]^, where HCW identifying as female were less likely to accept the COVID-19 vaccine as compared to male HCWs, our study found female HCWs to be more accepting of vaccination. Furthermore, similar to the findings of their study, we found HCWs in direct patient care to be more accepting of vaccination against COVID-19 compared to those HCWs who were involved in indirect patient care. Acceptability by age in our study was higher among the 51-60 years’ group, which is similar to other studies where increasing age and education are both positive factors in vaccine acceptance.^[17,24,25]^

To date, few studies have investigated acceptance of COVID-19 vaccines specifically among HCWs. A survey from France demonstrated a positive response for vaccine acceptance among majority of the participants (71.2%) and outright refusal was associated with female gender, age, lower educational status, and no report of chronic disease. ^[11]^ In contrast to these results, female gender was more likely to get vaccinated in our study and percentage of vaccine acceptance increased with increasing age.

Being a predominantly Muslim country, religion has often been a strong factor in rejecting vaccination for various vaccine preventable diseases in Pakistan, with many citing the contents of the vaccines to be non-compliant to Sharia law and therefore religiously unacceptable to them. ^[12,13,26,27]^ These findings were also reflected in our study, even among highly educated HCWs, particularly those who were female. However, recent public statements by major Islamic organizations have outlined that no such incompatibility exits ^[14]^.

Although, not directly addressed in our study, one important aspect of COVID-19 vaccine hesitancy in Pakistan, was the impact of social media as a source of information for HCWs during this pandemic. Social media posts have been implicated in similar studies carried out in Muslim majority Middle Eastern countries.^[17,28]^ Combatting this ‘infodemic’ with timely, evidence based communication is necessary to ensure that misinformation does not hamper national vaccination efforts^[29]^.

A major strength of our study was the robust sample size of HCWs who responded to the questionnaire and this survey represents a diverse group of individuals working as health care providers. However, there were some limitations to this study. A snowball sampling method could have created a selection and social desirability bias among HCWs. Furthermore, English questionnaire can produce a selection bias towards English-literate HCWs, particularly those active on social media. Despite these limitations, an overall positive response to vaccine acceptability is a positive sign towards attaining herd immunity worldwide, and increasing information and health communication from healthcare workers to the general population will decrease hesitancy towards COVID-19 vaccines.

## Conclusion

In conclusion, this survey suggests that early on in a vaccination drive, majority of the HCWs in Pakistan are willing to be vaccinated and only a small number of participants would actually reject being vaccinated. Overall, we gathered positive response towards COVID-19 vaccines but specific concerns regarding effectiveness and side effects were prevalent. Differences in vaccine acceptance were seen in various health care specialties, age groups, and ethnicity. Acceptance of COVID-19 vaccine in Pakistan is influenced by the evidence of vaccine effectiveness and while the acceptability among HCWs in Pakistan is higher than other surveys, a clear communication by the government, using the experience of HCWs as trusted sources of medical information, is needed to ensure the success of a national vaccination strategy.

## Data Availability

Available upon request

## Data availability

All the data presented in this manuscript is the copyright of Foundation University and there is legal restriction for sharing data publically by the Foundation University Ethical Review Committee (Number: FFH/51/DCA/2020). All HCW’s and other staff working in the University come under the employment of Fauji Foundation Head Quarters which abides by non-sharing of any anonymized or non-anonymized information about their employees. Data can be requested by contacting the Foundation University Ethical Review Committee at

## References

1. WHO WHO. Novel Coronavirus (2019-nCoV) SITUATION REPORT - 1 21 JANUARY 2020 [Internet]. Available from: https://www.who.int/docs/default-source/coronaviruse/situation-reports/20200121-sitrep-1-2019-ncov.pdf?sfvrsn=20a99c10_4

2. Huang C, Wang Y, Li X, Ren L, Zhao J, Hu Y, et al. Clinical features of patients infected with 2019 novel coronavirus in Wuhan, China. Lancet. 2020 Feb 15;395(10223):497–506. doi: 10.1016/S0140-6736(20)30183-5. Epub 2020 Jan 24. Erratum in: Lancet. 2020 Jan 30;: PMID: 31986264; PMCID: PMC7159299.

3. Rodriguez-Morales AJ, Cardona-Ospina JA, Gutiérrez-Ocampo E, Villamizar-Peña R, Holguin-Rivera Y, Escalera-Antezana JP, et al. Clinical, laboratory and imaging features of COVID-19: A systematic review and meta-analysis. Travel Med Infect Dis. 2020 Mar-Apr;34:101623. doi: 10.1016/j.tmaid.2020.101623. Epub 2020 Mar 13. PMID: 32179124; PMCID: PMC7102608.

4. WHO Coronavirus Disease (COVID-19) Dashboard | WHO Coronavirus Disease (COVID-19) Dashboard [Internet]. Available from: https://covid19.who.int/

5. Center NCO. National Command Operation Center [Internet]. Available from: https://ncoc.gov.pk/

6. Javed N, Khawaja H, Malik J, Ahmed Z. Endocrine dysfunction in psychology during social distancing measures. Bratisl Lek Listy 2020;121(12):878–80.

7. Lucia VC, Kelekar A, Afonso NM. COVID-19 vaccine hesitancy among medical students. J Public Health (Oxf). 2020 Dec 26:fdaa230. doi: 10.1093/pubmed/fdaa230. Epub ahead of print. PMID: 33367857; PMCID: PMC7799040.

8. Pakistan Starts COVID-19 Inoculation Drive | Voice of America - English [Internet]. Available from: https://www.voanews.com/covid-19-pandemic/pakistan-starts-covid-19-inoculation-drive

9. Coustasse A, Kimble C, Maxik K. COVID-19 and Vaccine Hesitancy: A Challenge the United States Must Overcome. J Ambul Care Manage. 2021 Jan/Mar;44(1):71–75. doi: 10.1097/JAC.0000000000000360. PMID: 33165121.

10. Fisher KA, Bloomstone SJ, Walder J, Crawford S, Fouayzi H, Mazor KM. Attitudes Toward a Potential SARS-CoV-2 Vaccine: A Survey of U.S. Adults. Ann Intern Med. 2020 Dec 15;173(12):964–973. doi: 10.7326/M20-3569. Epub 2020 Sep 4. PMID: 32886525; PMCID: PMC7505019.

11. Schwarzinger M, Watson V, Arwidson P, Alla F, Luchini S. COVID-19 vaccine hesitancy in a representative working-age population in France: a survey experiment based on vaccine characteristics. Lancet Public Health. 2021 Apr;6(4):e210–e221. doi: 10.1016/S2468-2667(21)00012-8. Epub 2021 Feb 6. PMID: 33556325; PMCID: PMC7864787.

12. Saied SM, Saied EM, Kabbash IA, Abdo SAE. Vaccine hesitancy: Beliefs and barriers associated with COVID-19 vaccination among Egyptian medical students. J Med Virol. 2021 Feb 28:10.1002/jmv.26910. doi: 10.1002/jmv.26910. Epub ahead of print. PMID: 33644891; PMCID: PMC8013865.

13. Robertson E, Reeve KS, Niedzwiedz CL, Moore J, Blake M, Green M, Katikireddi SV, Benzeval MJ. Predictors of COVID-19 vaccine hesitancy in the UK household longitudinal study. Brain Behav Immun. 2021 May;94:41–50. doi: 10.1016/j.bbi.2021.03.008. Epub 2021 Mar 11. PMID: 33713824; PMCID: PMC7946541.

14. Green MS, Abdullah R, Vered S, Nitzan D. A study of ethnic, gender and educational differences in attitudes toward COVID-19 vaccines in Israel - implications for vaccination implementation policies. Isr J Health Policy Res. 2021 Mar 19;10(1):26. doi: 10.1186/s13584-021-00458-w. PMID: 33741063; PMCID: PMC7977502.

15. Ali M, Khan DM, Aamir M, Khalil U, Khan Z. Forecasting COVID-19 in Pakistan. PLoS One. 2020 Nov 30;15(11):e0242762. doi: 10.1371/journal.pone.0242762. PMID: 33253248; PMCID: PMC7703963.

16. Butler R, MacDonald NE. Diagnosing the determinants of vaccine hesitancy in specific subgroups: The Guide to Tailoring Immunization Programmes (TIP). Vaccine [Internet] 2015;33(34):4176–9. Available from: https://linkinghub.elsevier.com/retrieve/pii/S0264410X15005022

17. Elbarazi I, Al-Hamad S, Alfalasi S, Aldhaheri R, Dubé E, Alsuwaidi AR. Exploring vaccine hesitancy among healthcare providers in the United Arab Emirates: a qualitative study. Hum Vaccines Immunother 2020;1–8.

18. Harapan H, Wagner AL, Yufika A, Winardi W, Anwar S, Gan AK, et al. Acceptance of a COVID-19 Vaccine in Southeast Asia: A Cross-Sectional Study in Indonesia. Front Public Health [Internet] 2020;8:381. Available from: https://www.frontiersin.org/article/10.3389/fpubh.2020.00381/full

19. Kabamba Nzaji M, Kabamba Ngombe L, Ngoie Mwamba G, Banza Ndala DB, Mbidi Miema J, Luhata Lungoyo C, et al. Acceptability of Vaccination Against COVID-19 Among Healthcare Workers in the Democratic Republic of the Congo. Pragmatic Obs Res [Internet] 2020;Volume 11:103–9. Available from: https://www.dovepress.com/acceptability-of-vaccination-against-covid-19-among-healthcare-workers-peer-reviewed-article-POR

20. Abid K, Bari YA, Younas M, Tahir Javaid S, Imran A. Progress of COVID-19 Epidemic in Pakistan. Asia Pac J Public Health [Internet] 2020;32(4):154–6. Available from: http://journals.sagepub.com/doi/10.1177/1010539520927259

21. Reinfection of COVID-19 in Pakistan: A First Case Report [Internet].;Available from: https://www.ncbi.nlm.nih.gov/pmc/articles/PMC7689968/

22. Lopez-Leon S, Wegman-Ostrosky T, Perelman C, Sepulveda R, Rebolledo PA, Cuapio A, et al. More than 50 Long-term effects of COVID-19: a systematic review and meta-analysis. medRxiv [Internet] 2021;Available from: https://www.ncbi.nlm.nih.gov/pmc/articles/PMC7852236/

23. Shekhar R, Sheikh AB, Upadhyay S, Singh M, Kottewar S, Mir H, et al. COVID-19 Vaccine Acceptance among Health Care Workers in the United States. Vaccines [Internet] 2021;9(2):119. Available from: https://www.mdpi.com/2076-393X/9/2/119

24. Karafillakis E, Dinca I, Apfel F, Cecconi S, Würz A, Takacs J, et al. Vaccine hesitancy among healthcare workers in Europe: A qualitative study. Vaccine 2016;34(41):5013–20.

25. Sallam M. COVID-19 vaccine hesitancy worldwide: a systematic review of vaccine acceptance rates:34.

26. Murakami H, Kobayashi M, Hachiya M, Khan ZS, Hassan SQ, Sakurada S. Refusal of oral polio vaccine in northwestern Pakistan: a qualitative and quantitative study. Vaccine 2014;32(12):1382–7.

27. Khattak FA, Rehman K, Shahzad M, Arif N, Ullah N, Kibria Z, et al. Prevalence of Parental refusal rate and its associated factors in routine immunization by using WHO Vaccine Hesitancy tool: A Cross sectional study at district Bannu, KP, Pakistan. Int J Infect Dis IJID Off Publ Int Soc Infect Dis 2020;104:117–24.

28. Sallam M, Dababseh D, Eid H, Al-Mahzoum K, Al-Haidar A, Taim D, et al. High Rates of COVID-19 Vaccine Hesitancy and Its Association with Conspiracy Beliefs: A Study in Jordan and Kuwait among Other Arab Countries. Vaccines [Internet] 2021;9(1). Available from: https://www.ncbi.nlm.nih.gov/pmc/articles/PMC7826844/

29. Infodemic [Internet].;Available from: https://www.who.int/health-topics/infodemic

